# The Impact of Pulmonary Fibrosis on Sex and Sexual Function – A Multinational Mixed Methods Study

**DOI:** 10.1101/2024.09.29.24314583

**Authors:** Na’ama Avitzur, Madelyn Knaub, Francesca Thornton-Wood, Simon R. Johnson, Christopher J. Ryerson, R. Gísli Jenkins, Iain Stewart, Kerri A. Johannson

## Abstract

**Background:** Sex is an important part of life for many adults, yet sexual function may be impacted by chronic respiratory diseases such as pulmonary fibrosis (PF). This multinational study sought to characterize the impact of PF on sex and sexual function, using mixed quantitative and qualitative methodology.

**Methods:** Patients were retrospectively included from a prospective registry and prospective clinical cohort if they had completed UCSD-SOBQ or SPARC questionnaire, respectively. An online multi-lingual survey used the Changes in Sexual Function Questionnaire (CSFQ) to assess sexual dysfunction, and qualitative evaluation of individual patient interviews was conducted using thematic analysis.

**Results:** Dyspnea with sexual activity affected 2,054/2,759 (74%) of registry patients, associated with male sex, lower FVC%, lower DLCO%, and worse cough. Distress due to the effect of PF on their sex life was reported in 52/225 (23%) of the clinical cohort, associated with younger age, male sex, lower DLCO%, and worse cough. Sexual dysfunction was common, affecting 56/67 (83%) of female and 63/73 (86%) male survey respondents. Qualitative analysis of patient interviews identified several themes including sex life limitations, changes in inter-personal relationships, quality of life, and emotions. All patients wanted to discuss sex with trusted healthcare providers.

**Conclusion:** In this multinational study, patients with PF reported engaging in sex and sexual activities but were adversely impacted by the effect of PF on sex life, with both physical and psychological limitations. Sexual dysfunction was common, driven by multiple disease domains. Sexual health appears to be an important component of comprehensive patient care.

**Funding:** The Canadian Registry for Pulmonary Fibrosis is sponsored by Boehringer Ingelheim, but had no input on any aspect of this study.

## Introduction

Pulmonary fibrosis (PF) includes a large group of lung diseases characterized by inflammation and fibrosis of the lung parenchyma. Many forms of PF are progressive and associated with high symptom burden, major morbidity, and early mortality; to date there is no cure for PF. PF most commonly affects adults, with increasing incidence and prevalence with older age, and relatively equal distribution across sexes and gender. Patients with PF frequently experience exercise limitation and symptoms such as exertional dyspnea, cough, and fatigue.^1,2^ PF is also associated with high comorbidity burden including cardiovascular disease, metabolic syndrome, multi-organ fibrosis, and the negative psychological impacts of depression and anxiety. All these factors result in patients with PF experiencing reduced overall and health-related quality of life.^3^

Sex and sexual function are important components of quality of life for many adults.^4^ Sexual function encompasses multiple components such as desire, arousal, orgasm, and satisfaction, among others. Sexual health is also influenced by psychological health, medications, and comorbidities. Large population-based studies have shown that most older adults engage in sex, with sexual dysfunction affecting 33-50%.^4,5^ Patients with respiratory disease experience symptoms and consequences that could affect sexual function, yet, little is known about sexual dysfunction in pulmonary conditions. Small studies suggest that sexual dysfunction is common in patients with pulmonary hypertension^6^ and COPD.^7,8^ In PF, most studies have focused on physical consequences of erectile dysfunction in men, or pelvic floor dysfunction in women, without assessing emotional wellbeing.^9–11^ This area warrants study, as PF patients identify sex and sexual function as a priority for research and assessment in clinical care.^3,12^

The objective of this multinational study was to characterize the impact of PF on sex and sexual function in a large group of affected adult patients, using mixed quantitative and qualitative methodology. We further sought to characterise associations of sexual dysfunction with other components of PF such as disease severity and oxygen use, and to understand the experience and needs of patients living with PF.

## Methods

The study used a mixed-methods approach with three components: retrospective analyses of a prospective multi-site registry and a prospective multi-site clinical study, a prospective multi-lingual online survey, and qualitative evaluation of patient interviews.

### Study populations

Patients with fibrotic ILD enrolled in the multi-center Canadian Registry for Pulmonary Fibrosis (CARE-PF) were included if they completed the University of California, San Diego Shortness of Breath Questionnaire (UCSD-SOBQ) at enrolment.^13^ Details of the CARE-PF cohort were previously described.^14^ Included patients had multidisciplinary diagnoses of idiopathic pulmonary fibrosis (IPF), connective tissue disease-related ILD (CTD-ILD), fibrotic hypersensitivity pneumonitis (fHP), or unclassifiable ILD. Age, sex, smoking history, contemporaneous (within 1 year) forced vital capacity percent predicted (FVC%) and diffusion capacity of the lung for carbon monoxide percent predicted (DLCO%), and patient recorded outcome measures including cough visual analogue scale (VAS)^15^ were extracted. Participants from the Prospective Observation of Fibrosis in Lung Clinical Endpoints (PROFILE) Central England cohort were included if they completed the Sheffield Profile for Assessment and Referral to Care (SPARC) questionnaire as part of a sub-study.^16^ The PROFILE cohort has been previously described and includes participants diagnosed with IPF and idiopathic fibrotic non-specific interstitial pneumonia (iNSIP)^17^. Baseline demographics, patient reported outcome measures including St George’s Research Questionnaire (SGRQ)^18^, Leicester Cough Questionnaire (LCQ)^19^, and lung function were ascertained.

The online survey was distributed through patient support/advocacy group email lists, newsletters, and social media. The online survey was available in four languages: English, French, Dutch, and Spanish, and was disseminated internationally to maximize representation and participation. Patients were invited to participate if they were adults (aged >/=18 years) with self-reported PF and able to complete the survey in one of the four available languages. The survey was anonymous with no identifying information collected, but participants could provide their email address if willing to be contacted for subsequent individual interviews.

### Exposure and outcome measures

Patients enrolled in CARE-PF completed the UCSD-SOBQ, a validated 24-item questionnaire that quantifies dyspnea in patients with respiratory disease. 21 questions enquired about dyspnea during specific activities with options to choose 0 (not breathless at all) to 5 (maximally breathless or too breathless to perform). Question 21 asked “When I do, or if I were to do, sexual activities, I would rate my shortness of breath as …” with scoring options from 0-5. Patients enrolled in PROFILE Central England Cohort completed SPARC, a 45-item tool developed to identify palliative care needs, and to distinguish a restricted set of items that could inform prognosis and clinical decisions in IPF, according to patient-reported distress.^16,17^ Question 31 asked “In the past month, have you been distressed or bothered by the effect of your condition on your sexual life?” with responses of 0 (not at all) to 3 (very much).

The online survey (**supplement**) collected demographics, type of PF and severity, including oxygen use and mMRC dyspnea score, and question 21 from UCSD-SOBQ and question 31 from SPARC.^20,21^ The remainder was based on the Changes in Sexual Functioning Questionnaire (CSFQ) – a sexual function survey validated in multiple languages for males and females, with a threshold score for sexual dysfunction.^22^ Participants were provided an introductory paragraph as per the CSFQ protocol (**supplement**), defining sexual activity as sexual intercourse, masturbation, sexual fantasies, and other sexual activities. The CSFQ assesses sexual desire, pleasure, arousal, and orgasm. This questionnaire was selected due to ease of distribution with only 14 questions, ability to assess both sexes with the same questionnaire, and because the questions were intended to be non-intrusive to facilitate survey uptake. Participants had the option to select non-binary gender, and, if selected, were asked to answer the male or female version based on their sexual organs. Analysis included participants who completed all survey questions. Sexual dysfunction scores were calculated using the sum of 14 CSFQ answers for a total score, with cut-off criteria as per the CSFQ protocol (<47 for males, and <41 for females).

### Qualitative Interviews and thematic analysis

Participants who provided their email address were contacted by one team member (NA) for participation in an optional 30-minute, one-on-one virtual interview. Interviews were conducted in English only, thus all participants needed verbal fluency in the English language. Patients did not identify themselves by name in the interview, and all participants agreed to recording and transcription using ZOOM™. NA conducted all interviews using an *a priori* structured guide of open-ended questions and prompts (**supplement**). Interviews were transcribed verbatim using ZOOM and edited by the interviewer (NA) for any discrepancies. Recruitment aimed for 10-15 participants or until theme saturation was achieved. Analysis was performed using the methodological principles of grounded theory.^23^ Two investigators (NA, MK) analysed and coded transcripts independently, using NVivo (version 14) software. Discrepancies were resolved by a third member of the study team (KJ). After analysing seven interviews independently, a codebook was developed and reviewed by NA and MK. Theme saturation was felt to be achieved by all three team members following seven interviews. Transcripts were coded a second time using the codebook, with agreement established. Direct quotes were extracted to present the themes.

### Statistical analysis

Study populations in registries and survey were characterised using descriptive statistics including means (SD) or medians (IQR) where appropriate. UCSD-SOBQ and SPARC scores were evaluated as binary (none vs any dyspnea or distress, respectively). Chi-square, t-test or Wilcox rank-sum was performed to test the relationship between presence of sexual dysfunction and categorical variables (gender, PF subtype, smoking status, oxygen desaturation, and dyspnea severity), normally distributed variables (FVC%, DLCO%, age) and non-normally distributed variables (UCSD total scores, SGRQ, LCQ, Cough VAS), respectively. Ordinal logistic regression evaluated associations between dyspnea scores and age, sex, lung function, and cough VAS in CARE-PF participants. Logistic regression was used to model PROFILE participants according to the binary outcome of no distress vs. any distress. Models were adjusted for lung function, age, sex/gender, lung function, and diagnosis. Models were specified separately in CARE-PF and PROFILE cohorts.

## Results

### Dyspnea with Sexual Activity

A total of 2,759 CARE-PF patients were included, with 1,119 (41%) having CTD-ILD, 866 (31%) with IPF, 515 (19%) with unclassifiable ILD, and 259 (9%) with fHP (**Table 1)**. A minority of patients (705, 26%) reported no dyspnea with sex (score=0), while 2,054 (74%) patients reported dyspnea with sexual activity (score>0). The median score for Q21 was 2 (IQR 0-4), with score distribution presented in **Figure 1a**. Dyspnea was more common in men (OR=1.2, 95%CI 1.1 to 1.4; p=0.009), in those who use oxygen (p<0.01), those with lower FVC% (OR=0.97, 95%CI 0.97 to 0.97; p<0.001), lower DLCO% (OR=0.98, 95%CI 0.97 to 0.98; p<0.001), and with higher cough VAS (OR=1.02, 95%CI 1.02 to 1.02; p<0.001). Self-reported dyspnea during sexual activity was not associated with age, or PF subtype **(Table 2)**. Probabilities of response of 0 or 5 for Q21 across FVC% and DLCO% are presented in **Figure 2**.

**Table 1.** CARE-PF and PROFILE patient demographics by exposure vs none.

| Cohort | CARE-PF (Canada) |  |  | PROFILE (UK) |  |  |
| --- | --- | --- | --- | --- | --- | --- |
| Parameter | Dyspnea with sexual activity (n=2054) | No dyspnea with sexual activity (n=705) | P-value | Distress due to effect of PF on sex life (n=52) | No distress due to effect of PF on sex life (n=173) | P-value |
| **Age (years)** | 64 ± 12 | 67 ± 11 | **p<0.001** | 67 ± 10 | 73 ± 8 | **p<0.001** |
| **Sex** |  |  | **p<0.001** |  |  | **p=0.014** |
| Female | 901 (44%) | 381 (54%) |  | 6 (12%) | 49 (28%) |  |
| Male | 1152 (56%) | 323 (46%) |  | 46 (88%) | 124 (72%) |  |
| **Oxygen users, %** | 568 (30%) | 58 (9%) | **p<0.001** | - | - | - |
| **Oxygen desaturation, %** | 539 (75%) | 153 (63%) | **p=0.001** | 8 (20%) | 26 (23%) | p=0.675 |
| **Type of PF, %** |  |  | p=0.739 |  |  | p=0.769 |
| IPF | 635 (31%) | 231 (33%) |  | 43 (83%) | 146 (84%) |  |
| CTD-ILD | 833 (41%) | 286 (40%) |  | - | - |  |
| Fibrotic HP | 195 (9%) | 64 (9%) |  | - | - |  |
| Unclassifiable ILD | 391 (19%) | 124 (18%) |  | - | - |  |
| Idiopathic NSIP | - | - | - | 9 (17%) | 27 (16%) |  |
| **UCSD-SOBQ score** | 45.4 ± 26 | 15.7 ± 16 | **p<0.001** | - | - | - |
| **FVC%** | 72.4 ± 20 | 82.4 ± 19 | **p<0.001** | 79.5 ± 20 | 82.8 ± 19 | p=0.281 |
| **DLCO%** | 55.3 ± 19 | 62.7 ± 19 | **p<0.001** | 43.0 ± 15 | 50.6 ± 16 | **p=0.006** |
| **Cough VAS** | 40.8 ± 24 | 31.0 ± 23 | **p<0.001** | - | - | - |
| **LCQ cough score** | - | - | - | 14.0 ± 4 | 16.6 ± 4 | **p<0.001** |

PF: pulmonary fibrosis, IPF: idiopathic pulmonary fibrosis, CTD-ILD: connective tissue disease - interstitial lung disease, HP: hypersensitivity pneumonitis, NSIP: non-specific interstitial pneumonia, UCSD-SOBQ: University of California, San Diego Shortness of Breath Questionnaire, FVC: forced vital capacity, DLCO: diffusion capacity of the lung for carbon monoxide, VAS: visual analogue score, LCQ: Leicester Cough Questionnaire Dyspnea based on answer ≥ 1 on UCSD-SOBQ Q#21. Distress based on answer ≥ 1 on SPARC Q#31.
\*Percentages based on non-missing data.

**Figure 1:**
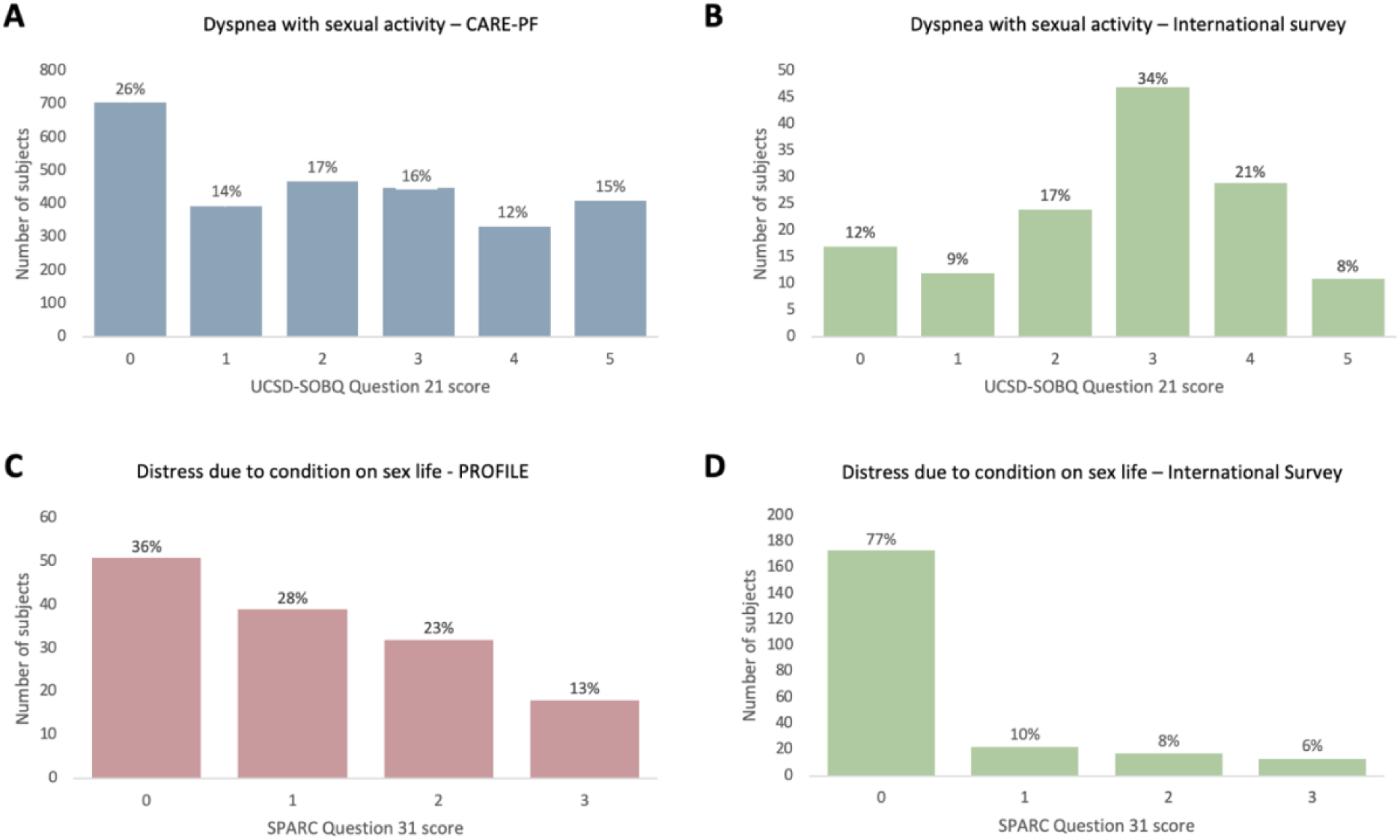
Distribution of participant answers to UCSD-SOBQ question #21 in CARE-PF (A) and international survey (B), SPARC question #31 in PROFILE (C) and international survey (D)

**Table 2.**
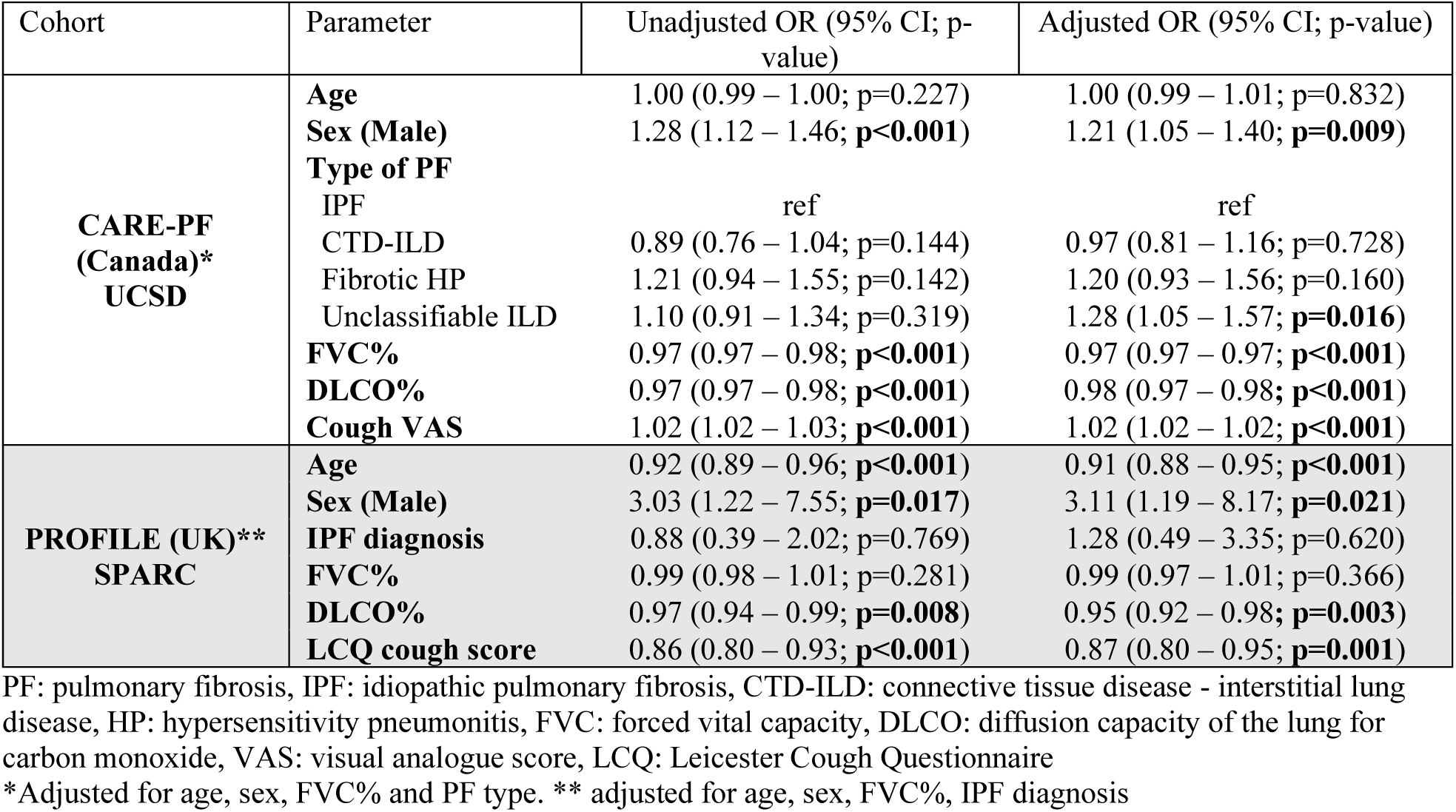
Logistic regression for UCSD-SOBQ Q#21, and SPARC Q#31 and clinical measures.

**Figure 2:**
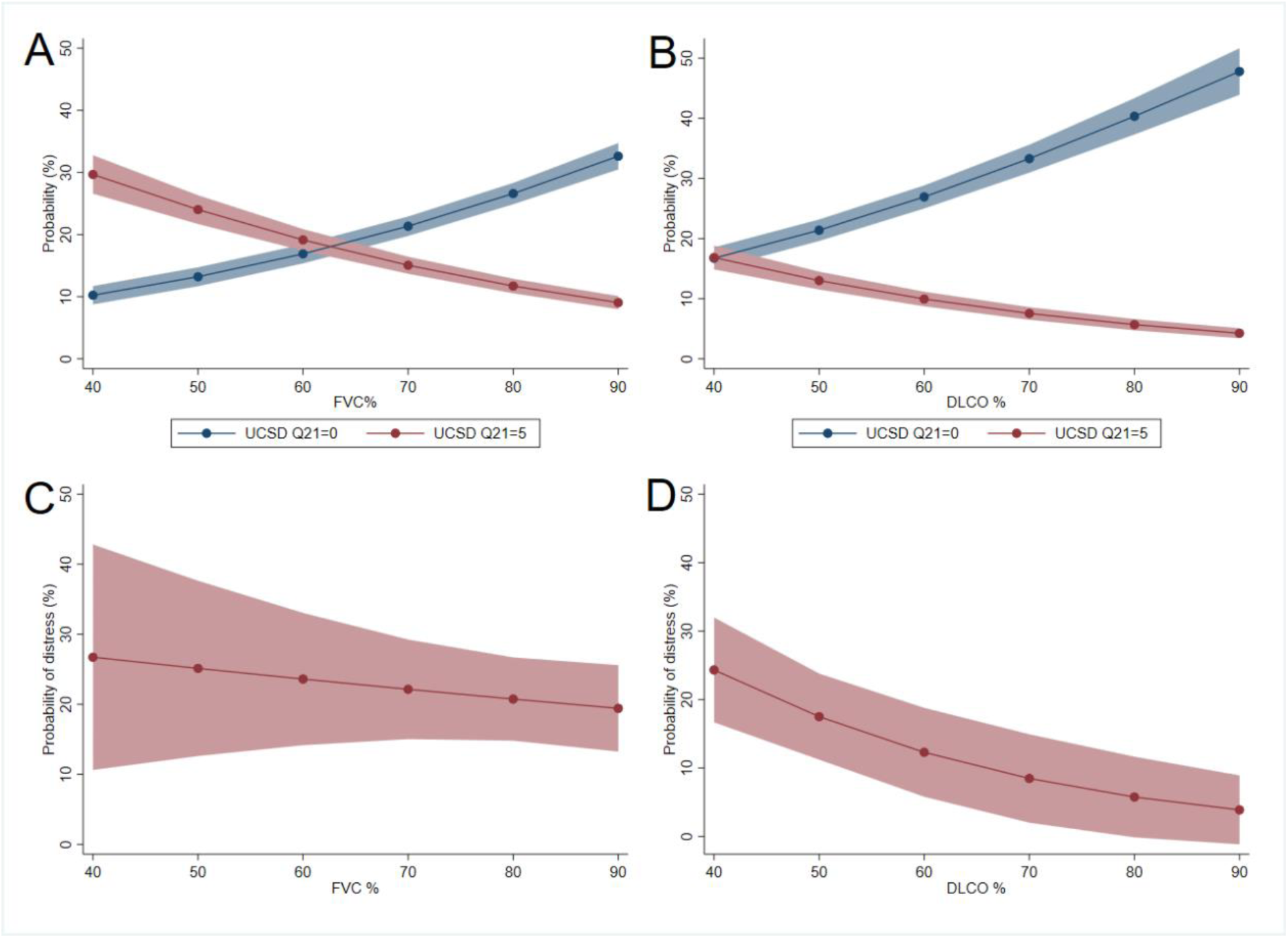
Probability of dyspnea relative to FVC % (A) and DLCO % (B). Probability of distress due to effect of patient condition on sex life relative to FVC % (C) and DLCO % (D). Models adjusted for age, sex, diagnosis.

### Distress due to effect of PF on sex life

A total of 225 PROFILE participants had complete SPARC responses, of which 189 had a diagnosis of IPF (84%) and 36 (16%) with fNSIP (**Table 1)**. Most patients (173, 77%) reported no distress in the past month due to the effect of PF on their sex life (score 0), while 52 (23%) reported some level of distress (score>0) (**Fig 1c)**. No difference was observed in distress according to IPF and iNSIP diagnosis. Distress was more common with younger age (OR=0.91, 95%CI 0.88 to 0.95; p<0.001), male sex (OR=3.11, 95%CI 1.19 to 8.17; p=0.02), lower DLCO% (OR=0.95, 95%CI 0.92 to 0.98; p=0.003), and lower LCQ indicating worse cough (OR=0.87, 95%CI 0.80 to 0.95; p<0.001). There was no association with FVC% (**Table 2**). Probability of reporting any distress across FVC% and DLCO% is presented in **Figure 2**.

### Online International Survey

The survey was disseminated broadly online, thus response rate cannot be determined. The survey was completed by 140 participants; 64 in English, 37 in Dutch, 31 in Spanish, and 8 in French (**Table 3**). Survey participants were younger than the CARE-PF and PROFILE cohorts, and a higher percentage used oxygen. Type of PF was predominantly IPF, followed by CTD-ILD. Median score for UCSD-SOBQ Q21 was 3 (IQR 2-4), and for SPARC Q31 was 1 (IQR 0-2), which were higher compared with the registry scores (**Figure 1b,d)**. Mean CSFQ score for females was 30 (±11), while for males it was 36 (±11). Sexual dysfunction was common, affecting 56/67 (83%) of females and 63/73 (86%) of males (**>Table 4**), consistent across all languages. Participants with sexual dysfunction were older (mean age 63.4 vs 52.7 years), but with lower mMRC dyspnea, compared with those without sexual dysfunction.

**Table 3.**
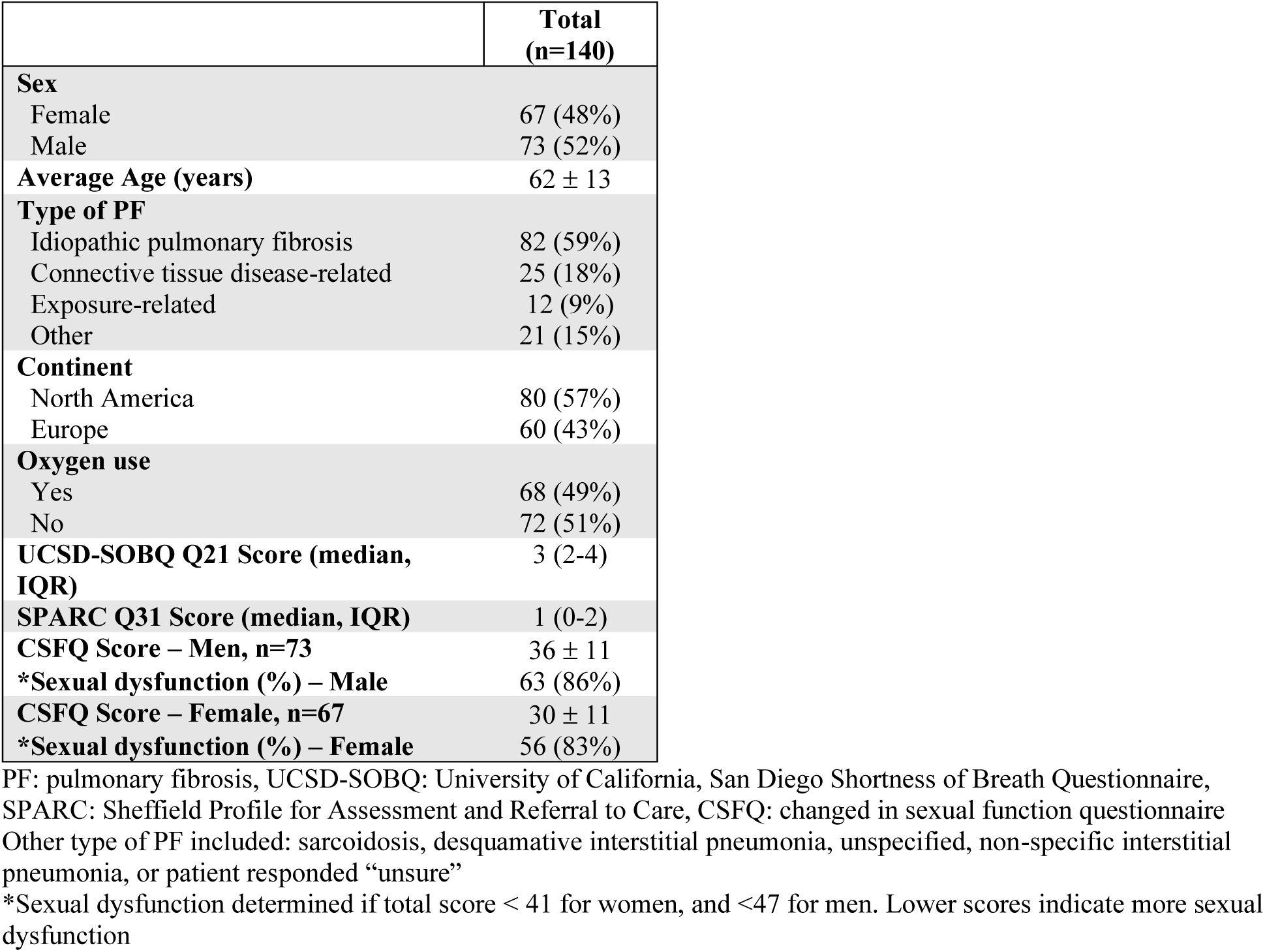
International survey results.

|  | **Total**<br>**(n=140)** |
| --- | --- |
| **Sex** |  |
| Female | 67 (48%) |
| Male | 73 (52%) |
| **Average Age (years)** | 62 ± 13 |
| **Type of PF** |  |
| Idiopathic pulmonary fibrosis | 82 (59%) |
| Connective tissue disease-related | 25 (18%) |
| Exposure-related | 12 (9%) |
| Other | 21 (15%) |
| **Continent** |  |
| North America | 80 (57%) |
| Europe | 60 (43%) |
| **Oxygen use** |  |
| Yes | 68 (49%) |
| No | 72 (51%) |
| **UCSD-SOBQ Q21 Score (median, IQR)** | 3 (2-4) |
| **SPARC Q31 Score (median, IQR)** | 1 (0-2) |
| **CSFQ Score – Men, n=73** | 36 ± 11 |
| **\*Sexual dysfunction (%) – Male** | 63 (86%) |
| **CSFQ Score – Female, n=67** | 30 ± 11 |
| **\*Sexual dysfunction (%) – Female** | 56 (83%) |

PF: pulmonary fibrosis, UCSD-SOBQ: University of California, San Diego Shortness of Breath Questionnaire, SPARC: Sheffield Profile for Assessment and Referral to Care, CSFQ: changed in sexual function questionnaire Other type of PF included: sarcoidosis, desquamative interstitial pneumonia, unspecified, non-specific interstitial pneumonia, or patient responded “unsure”
\*Sexual dysfunction determined if total score < 41 for women, and <47 for men. Lower scores indicate more sexual dysfunction

### Qualitative analysis

Seven interviews were completed. Due to small numbers and to maintain participant confidentiality, demographic information is not reported. Five major themes emerged throughout the interviews, including sex while living with pulmonary fibrosis, emotions, research, relationships, and quality of life (**Figure 3**). A full codebook can be found in the supplemental material.

**Figure 3:**
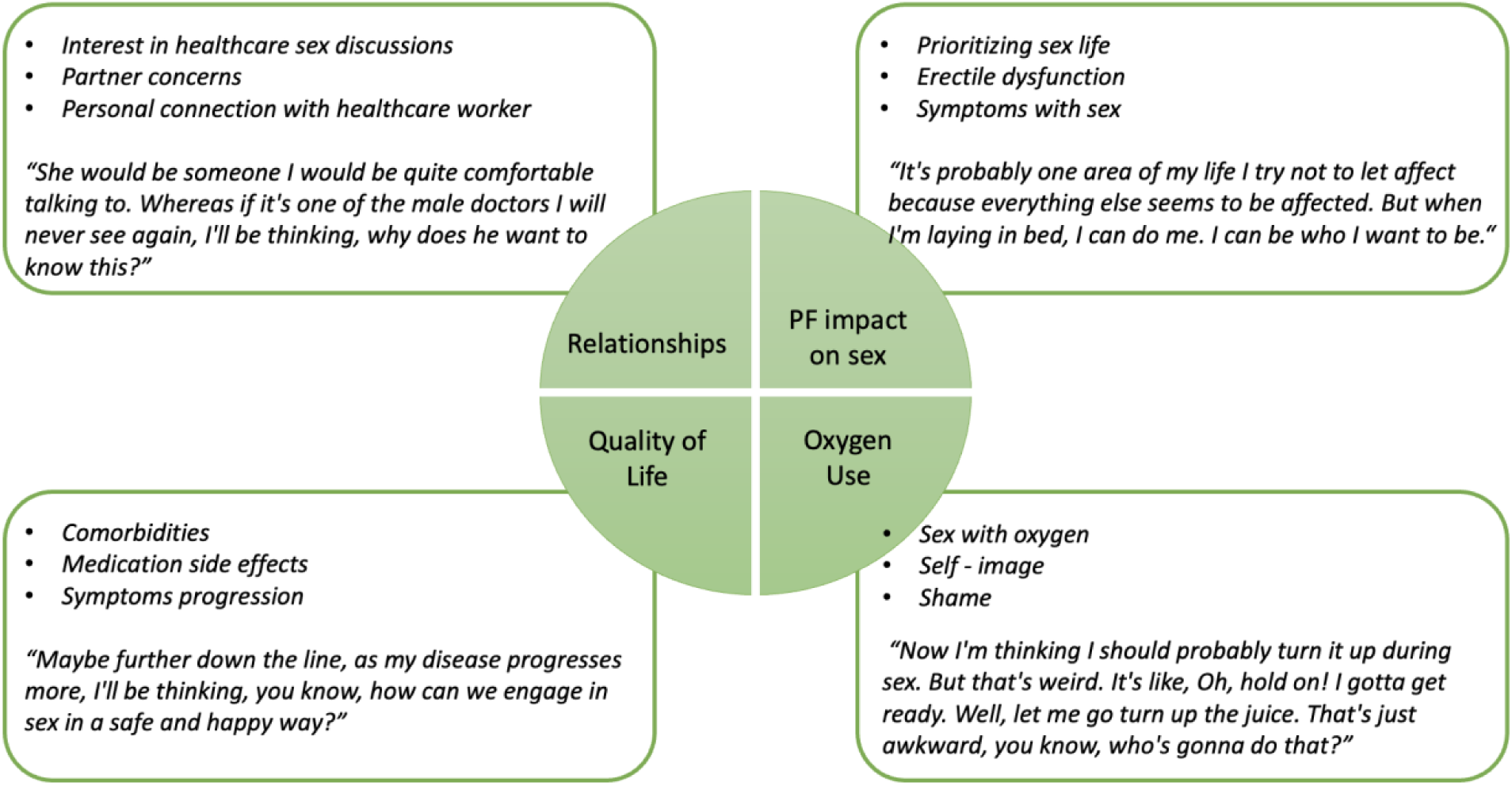
Qualitative analysis themes. Quotes were extracted from patient interviews.

#### Sex with pulmonary fibrosis

Participants discussed their current sex life, including symptoms with sex such as dyspnea, cough, and fatigue. All men experienced erectile dysfunction, often noted by them around the time of their PF diagnosis. Participants described prioritising their sex life, and how they have adapted in the context of PF. For example: *“It’s probably one area of my life I try not to let affect, because everything else seems to be affected. But when I’m laying in bed, I can do me. I can be who I want to be. I don’t have to worry about anything else you know, so it’s that big, my time”(participant #1).* Participants using oxygen described various challenges including cumbersomeness of using oxygen during sex, and oxygen influence on their self-image and shame (**Figure 3)**.

#### Emotions

Participants shared various perspectives around emotional states related to PF and their sex life, including worry and frustration around their condition, as well as frequent embarrassment, guilt, and shame in discussing sex as it related to their PF. One participant (#7) shared: “*I think the only thing that would limit me talking to a professional, is if I felt I was making them uncomfortable, or embarrassed. Then I would probably hold back in the discussion.”*

#### Relationships

Participants described the impact of PF and their sex life on inter-personal relationships. Two participants identified as LGBTQ+ and acknowledged that most research in PF does not include LGBTQ+ perspectives. Multiple participants had concerns around their sex life due to romantic relationship challenges, or their partner’s worry of injuring them during sex. All participants indicated they had never discussed sex with any healthcare provider, but they were all interested in discussing their sexual function, if they felt they had a strong relationship founded on trust. For example, participant #5 stated: “*All my doctors have seen the worst of me. The worst that my body can put me through. I’m very comfortable with all of them.”* Participants were comfortable having these discussions with their pulmonologist, family physician, nurse practitioner, or PF educator. One participant (#2) brought up considerations around timing of these conversation in the context of a new PF diagnosis: “*I think so, but not right away. You know I was scared to death when I got this diagnosis… So the last thing I’m thinking about is sex at that point.”*

#### Quality of life

Participants shared various consequences of PF that impact their quality of life, and therefore also impact their sex life. These included symptom progression, comorbidities, and medication side effects. Regarding antifibrotics one participant (#7) said: “*The treatment is a horrible little pill called [sic antifibrotic], and two of the side effects are vomiting and diarrhea. So there will be occasions…we may have felt in the mood for intimacy. The pill has decided that it’s going to make me violently sick. So you have to put your sexual activity on the sidelines and wait until that side of life passes.”*

Participants discussed participation in pulmonary rehabilitation, but only one reported a session on sex and intimacy as part of the curriculum.

#### Research

A recurring theme was participants’ desire and experience in engaging in research and learning around PF and sexual function. All participants valued the importance of research engagement, knowledge distribution, and the ability to find new research related to PF. Some commented that they would find further research on sex in PF valuable, particularly the link between erectile dysfunction and PF. Participants expressed enthusiasm around the possibility of improving patients’ sexual function and in turn, their quality of life.

## Discussion

In this multi-national mixed methods study, we demonstrated that PF impacts sex and sexual function of affected individuals across multiple domains. Patients with PF reported engaging in sexual activities, even in advanced stages of disease with low lung function and while requiring supplemental oxygen. Patients expressed desire to have resources for support and education in the context of their disease, and for future work to include sex as an important aspect of quality of life.

Sexual dysfunction has been studied in other lung diseases, including pulmonary hypertension and COPD.^6,8^ In PF patients, the first study in this area is from 1984, which studied 8 men with IPF, showing that erectile dysfunction was common, and reporting an association of low serum testosterone with hypoxemia^24^. Additional small studies in 90 women with systemic sclerosis, and 54 men with ILD reported high frequency of sexual dysfunction, and erectile dysfunction, respectively ^10,9^. A recent single centre study of 93 patients with sarcoidosis found that 67% of women had sexual dysfunction, and 37% of men had erectile dysfunction^11^. These studies have typically included one gender/sex, in a single centre with small samples sizes, limiting generalisability of findings, and focusing primarily on physiological aspects. Our findings build on these preliminary data with greater international generalisability, demonstrate associations with clinical outcomes, present patient voices and priorities, thus providing novel insights into sexual dysfunction in patients with chronic pulmonary disease. Importantly, this is the first study to have included the experience and voices of LGBTQ+ patients living with PF.

Both the Canadian and UK cohort data revealed that PF impacts patients’ symptoms with, and emotions around sex. Unsurprisingly, increased dyspnea with sexual activity was associated with lower lung function, and cough. However, and importantly, sex-associated patient distress was associated with lower DLCO% and increased cough. This suggests that disease severity and symptom burden negatively impact patients’ sex lives via physical domains and the emotional wellbeing associated with sex. Whether interventions that improve PF-related symptoms such as cough or lung function can translate into improved sexual health warrants further study.

We recruited a large number of patients to our survey, finding higher rates of sexual dysfunction among both males and females compared to other studies of adult populations. A United States-based study of 3000 community-dwelling older adults found that about 50% of sexually active adults had at least one bothersome symptom with sex.^4^ Additionally, sexual dysfunction can affect 18-60% of adults with chronic diseases such as cirrhosis or hematological malignancies.^25,26^ Thus, the 80% rate of sexual dysfunction in our population was much higher than that of the general population of older adults, and most likely due to underlying components of PF. Furthermore, our survey population was younger than those in the registries, suggesting sexual dysfunction affects both younger and older PF patients. We found similar rates of sexual dysfunction across languages and PF subtypes highlighting the impact across the PF population.

The qualitative analysis introduced essential themes that are not identifiable using quantitative data. In particular, we found that patients had limited or no discussions about sex with their healthcare providers, despite wanting such discussions. They were also interested in obtaining further information about sex and PF online or through programs. Despite various challenges associated with PF, such as medication side effects, oxygen use, and symptoms, all patients wanted to have an active sex life. One patient mentioned a pulmonary rehabilitation program that included a dedicated sex and intimacy session in the context of advanced lung disease, but this is not a mandatory component of pulmonary rehabilitation, and it is unknown how many programs specifically address this. Overall, sex was described as an important component of quality of life, and pulmonary rehabilitation programs may provide an optimal opportunity to address sexual health.

Similar to other studies we found high rates of erectile dysfunction (ED).^6,9^ Rates of ED are estimated at 40% in the general population,^4^ while our survey demonstrated close to 80%, and all male interviewees endorsed ED. Men were also more likely than woman to report dyspnea and distress. Recent data suggest that fibrotic diseases may be shared across multiple organs, including the reproductive organs and genitalia.^27^ Whether multimorbid and cross-organ fibrosis can account for these relationships between PF and sexual dysfunction warrants further investigation.

Based on our findings, we have developed a framework by which PF impacts sexual function, through the multi-faceted patient disease journey (**Figure 4**). This includes early features in diagnosis (symptoms, change in self-image), disease progression (physiologic function, medication side effects), end-of-life factors (oxygen use and emotional distress), and finally strain on romantic relationships. This conceptual framework can provide a structure around which to build opportunities for targeted intervention to optimise sexual wellbeing and promote research that improves patients’ quality of life.

**Figure 4:**
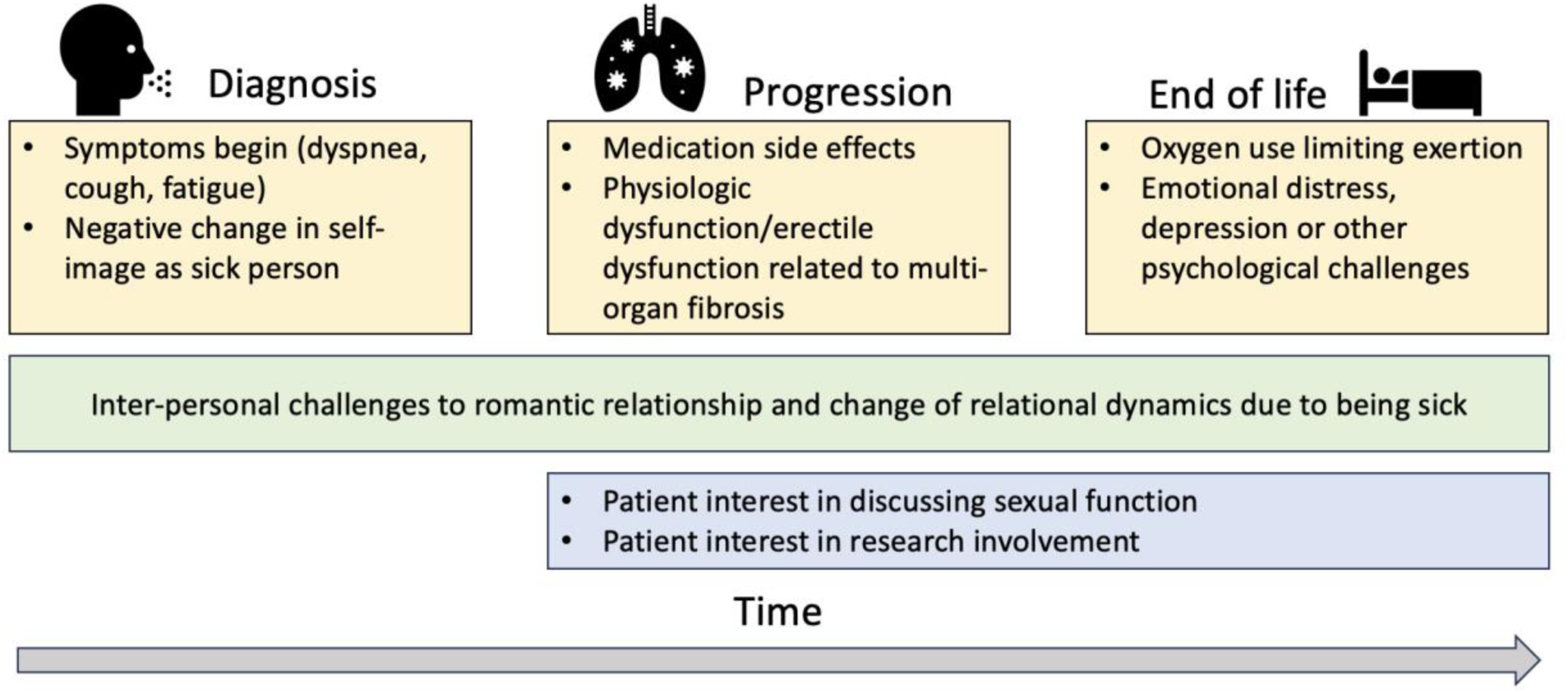
Key factors leading to sexual dysfunction throughout pulmonary fibrosis disease course. Early in diagnosis, symptoms limit sexual activity, and a patient’s self-image as a newly sick person impacts their sexual desire. Physiologic dysfunction becomes more common over time, and undesired medications side effects can lead to avoiding intimacy. At end of life, oxygen use again affects self-image, and carries practical challenges while engaging in sex. Throughout all phases of disease, patients can experience changes in their romantic relationships, often reducing sexual activity. Patients are interested in discussing sexual function with their healthcare team, but timing is preferred following the initial diagnosis.

This study has limitations. The patient cohort studies are from Canada and the UK, and the qualitative measures were available in English alone, however the survey was in multiple languages, mitigating missing representation of the general population. Survey and interview participants chose to participate, likely introducing selection bias, but other data was soured from prospective observational studies. Sexuality is a complex individual and social construct that likely differs in diverse regions of the world, and despite efforts to include diverse patients, not all experiences were studied. We could not control for underlying depression, anxiety, or other comorbidities which may impact sexual function. Not all forms of PF were represented, however our data suggest consistency in dyspnea and distress across the PF subtypes that were included. The broad range of questionnaire scores, oxygen usage rates, ages and disease severities supports that sex is an important issue across the disease spectrum, and may change as a patient’s disease progresses.

In summary, this is the largest and most comprehensive study of sex and sexual function in patients with PF including multiple subtypes, sexes, genders and sexual orientations. We have included multiple key voices and lived experiences to characterise this previously under-recognised issue. We have shown that sexual dysfunction is common in patients with PF, driven by multiple domains of disease, and not exercise limitation alone. These domains include symptoms, oxygen use, disease severity, medication side-effects, self-image, and relationships. Taken together, our findings highlight the importance of providing comprehensive care to patients with PF beyond physiologic measures of lung function, as well as the need to use standardised quality of life tools to measure sexual dysfunction in PF. Sex is an important aspect of patients’ quality of life, and further research is warranted to identify mechanisms underlying sexual dysfunction, and to identify interventions that optimise patients’ sexual well-being.

## Data Availability

Data produced in the present work are available upon reasonable request to the authors

## Acknowledgments

We would like to express our deep gratitude to all the patients who completed these surveys, questionnaires and interviews for sharing their experiences, and supporting this work. We hope we have represented your views holistically.

We would also like to gratefully acknowledge Dr. Maria Molina, Dr. Marlies Wijsenbeek, Dr. Catherina Moor, Dr. Pilar Rivera-Ortega and Dr. Sarah Hosseini for their invaluable assistance with translation of the surveys. We would like to acknowledge the British Association of Lung Research for their financial support of FTW, and the NIHR Nottingham Biomedical Research Centre.

Thank you to our national and international colleagues, the Canadian Pulmonary Fibrosis Foundation, and Action for Pulmonary Fibrosis for distributing the survey to patient groups. CARE-PF collaborators: Dr. Jolene Fisher, Dr. Martin Kolb, Dr. Veronica Marcoux, Dr. Hélène Manganas, Dr. Nasreen Khalil, Dr. Deborah Assayag.

## Contributions

NA, and KAJ participated in the design and planning of the study. All authors provided critical input on study design, data analysis and interpretation, and manuscript preparation.

