## Supplemental material for "The Impact of Pulmonary Fibrosis on Sex and Sexual Function – A Multinational Mixed Methods Study"

### **Online survey questions – English Version**

Length: 10 minutes

Thank you for participating in this survey. The purpose of this project is to understand how pulmonary fibrosis (PF) may impact patients’ sexual activity.

This is a questionnaire about sexual activity and sexual function. By sexual activity, we mean sexual intercourse, masturbation, sexual fantasies and other sexual activities.

Pulmonary fibrosis (PF) can affect every area of a patient’s life, including having an impact on sexual activity. We want to understand the changes PF may have on a person’s intimate sexual relationships. We will use this information to adapt care and help patients improve their quality of life.

This survey was designed by the University of Calgary’s Interstitial Lung Disease Program, and adapted from the Changes in Sexual Functioning Questionnaire. This survey is anonymous and all data will be confidential.

This study has been approved by the University of Calgary Conjoint Health Research Ethics Board (REB22-1570). Your participation is completely voluntary. The survey will ask you questions about PF and your thoughts on, and experience of your sexual function. You can withdraw from the survey at any time, simply close your internet browser and no data will be collected. We do not foresee any risks to patients or family members by participating in this project. The researchers have no conflicts of interests to declare, pertaining to this study. By completing this online survey, you confirm you are over the age of 18 and consent to have your answers used to better understand the issue of sexual function for PF.

If you have any questions or would like a copy of this form, please

Clicking the forward button will bring you to the consent form for this study. Clicking the back button will exit you from this study and end your participation.

1. Sex

1. Male
2. Female
3. Non-binary
4. Prefer not to answer

2. Age:

- Blank

3. What kind of interstitial lung disease do you have?

1. Idiopathic Pulmonary Fibrosis (IPF)
2. Connective tissue disease ILD (Rheumatoid arthritis, Scleroderma, Sjogren’s, Lupus etc.)
3. Exposure related (hypersensitivity pneumonitis, silicosis, asbestosis)
4. I am not sure
5. Other (Specify below)

4. Based on what you know, how severe is your pulmonary fibrosis (based on symptoms, breathing tests, and 6 minute walk test)?

1. Mild
2. Moderate
3. Severe
4. I am not sure

5. Do you use oxygen?

1. Yes
2. No

6. When do you become short of breath? Please choose the description that applies to you.

0. I only get breathless with strenuous exercise

1. I get short of breath when hurrying on flat ground, or walking up a slight hill
2. I walk slower than people my age on flat ground because of breathlessness
3. I have to stop and catch my breath after walking 100 yards (90m) or a few minutes on flat ground
4. I am breathless when getting dressed or I am too breathless to leave the house

7. When I do sexual activities, I would rate my shortness of breath as

0. None

1. Minimal

2. Mild

3. Moderate

4. Severe

5. Maximum or unable to do because of shortness of breath

8. In the past month, have you been distressed or bothered by the effect of your condition on your sexual life?

0. None at all

1. A little bit

2. Quite a bit

3. Very much

CSFQ for men or women based on answer to question #1 – see below both versions.

If respondent indicates “non-binary” or “prefer not to answer” in question #1, provide them a choice for male or female version based on their organs.

NOTE**: This is a questionnaire about sexual activity and sexual function. By sexual activity, we mean sexual intercourse, masturbation, sexual fantasies and other sexual activities.**

**Female Version**

1. How enjoyable or pleasurable is your sex life right now, compared with the most enjoyable it has ever been?

🞏 1-No enjoyment or pleasure

🞏 2-Little enjoyment or pleasure

🞏 3-Some enjoyment or pleasure

🞏 4-Much enjoyment or pleasure

🞏 5-Great enjoyment or pleasure

2. How often do you engage in sexual activities (sexual intercourse, masturbation, etc.) now?

🞏 1-Never

🞏 2-Rarely (once a month or less)

🞏 3-Sometimes (more than once a month, up to twice a week)

🞏 4-Often (more than twice a week)

🞏 5-Every day

3. How often do you want to engage in sexual activities?

🞏 1-Never

🞏 2-Rarely (once a month or less)

🞏 3-Sometimes (more than once a month, up to twice a week)

🞏 4-Often (more than twice a week)

🞏 5-Every day

4. How often do you have sexual thoughts (thinking about sex, sexual fantasies) now?

🞏 1-Never

🞏 2-Rarely (once a month or less)

🞏 3-Sometimes (more than once a month, up to twice a week)

🞏 4-Often (more than twice a week)

🞏 5-Every day

5. Do you enjoy books, magazines, movies, music or artwork with sexual or erotic content?

🞏 1-Never

🞏 2-Rarely (once a month or less)

🞏 3-Sometimes (more than once a month, up to twice a week)

🞏 4-Often (more than twice a week)

🞏 5-Every day

6. How much pleasure or enjoyment do you get from sexual thoughts and fantasies?

🞏 1-No enjoyment or pleasure

🞏 2-Little enjoyment or pleasure

🞏 3-Some enjoyment or pleasure

🞏 4-Much enjoyment or pleasure

🞏 5-Great enjoyment or pleasure

7. How often do you become sexually aroused?

🞏 1-Never

🞏 2-Rarely (once a month or less)

🞏 3-Sometimes (more than once a month, up to twice a week)

🞏 4-Often (more than twice a week)

🞏 5-Every day

8. Are you easily aroused?

🞏 1-Never

🞏 2-Rarely (much less than half the time)

🞏 3-Sometimes (about half the time)

🞏 4-Often (much more than half the time)

🞏 5-Always

9. Is your vaginal lubrication adequate during sexual activities?

🞏 1-Never

🞏 2-Rarely (much less than half the time)

🞏 3-Sometimes (about half the time)

🞏 4-Often (much more than half the time)

🞏 5-Always

10. How often do you become aroused and then lose interest during sexual activities?

🞏 5-Never

🞏 4-Rarely (much less than half the time)

🞏 3-Sometimes (about half the time)

🞏 2-Often (much more than half the time)

🞏 1-Always

11. How often do you have an orgasm?

🞏 1-Never

🞏 2-Rarely (much less than half the time)

🞏 3-Sometimes (about half the time)

🞏 4-Often (much more than half the time)

🞏 5-Always

12. Are you able to have an orgasm when you want to during sexual activities?

🞏 1-Never

🞏 2-Rarely (much less than half the time)

🞏 3-Sometimes (about half the time)

🞏 4-Often (much more than half the time)

🞏 5-Always

13. How much pleasure or enjoyment do you get from your orgasms?

🞏 1-No enjoyment or pleasure

🞏 2-Little enjoyment or pleasure

🞏 3-Some enjoyment or pleasure

🞏 4-Much enjoyment or pleasure

🞏 5-Great enjoyment or pleasure

14. How often do you have painful orgasms?

🞏 5-Never

🞏 4-Rarely (once a month or less)

🞏 3-Sometimes (more than once a month, up to twice a week)

🞏 2-Often (more than twice a week)

🞏 1-Every day

**Male Version**

1. How enjoyable or pleasurable is your sex life right now, compared with the most enjoyable it has ever been?

🞏 1-No enjoyment or pleasure

🞏 2-Little enjoyment or pleasure

🞏 3-Some enjoyment or pleasure

🞏 4-Much enjoyment or pleasure

🞏 5-Great enjoyment or pleasure

2. How often do you engage in sexual activities (sexual intercourse, masturbation, etc.) now?

🞏 1-Never

🞏 2-Rarely (once a month or less)

🞏 3-Sometimes (more than once a month, up to twice a week)

🞏 4-Often (more than twice a week)

🞏 5-Every day

3. How often do you want to engage in sexual activities?

🞏 1-Never

🞏 2-Rarely (once a month or less)

🞏 3-Sometimes (more than once a month, up to twice a week)

🞏 4-Often (more than twice a week)

🞏 5-Every day

4. How often do you have sexual thoughts (thinking about sex, sexual fantasies) now?

🞏 1-Never

🞏 2-Rarely (once a month or less)

🞏 3-Sometimes (more than once a month, up to twice a week)

🞏 4-Often (more than twice a week)

🞏 5-Every day

5. Do you enjoy books, magazines, movies, music or artwork with sexual or erotic content?

🞏 1-Never

🞏 2-Rarely (once a month or less)

🞏 3-Sometimes (more than once a month, up to twice a week)

🞏 4-Often (more than twice a week)

🞏 5-Every day

6. How much pleasure or enjoyment do you get from sexual thoughts and fantasies?

🞏 1-No enjoyment or pleasure

🞏 2-Little enjoyment or pleasure

🞏 3-Some enjoyment or pleasure

🞏 4-Much enjoyment or pleasure

🞏 5-Great enjoyment or pleasure

7. How often do you have an erection related or unrelated to sexual activities?

🞏 1-Never

🞏 2-Rarely (once a month or less)

🞏 3-Sometimes (more than once a month, up to twice a week)

🞏 4-Often (more than twice a week)

🞏 5-Every day

8. Do you get an erection easily?

🞏 1-Never

🞏 2-Rarely (much less than half the time)

🞏 3-Sometimes (about half the time)

🞏 4-Often (much more than half the time)

🞏 5-Always

9. Are you able to maintain an erection during sexual activities?

🞏 1-Never

🞏 2-Rarely (much less than half the time)

🞏 3-Sometimes (about half the time)

🞏 4-Often (much more than half the time)

🞏 5-Always

10. How often do you experience painful, prolonged erections during sexual activities?

🞏 5-Never

🞏 4-Rarely (once a month or less)

🞏 3-Sometimes (more than once a month, up to twice a week)

🞏 2-Often (more than twice a week)

🞏 1-Every day

11. How often do you ejaculate during sexual activities?

🞏 1-Never

🞏 2-Rarely (once a month or less)

🞏 3-Sometimes (more than once a month, up to twice a week)

🞏 4-Often (more than twice a week)

🞏 5-Every day

12. Are you able to ejaculate when you want to during sexual activities?

🞏 1-Never

🞏 2-Rarely (much less than half the time)

🞏 3-Sometimes (about half the time)

🞏 4-Often (much more than half the time)

🞏 5-Always

13. How much pleasure or enjoyment do you get from your orgasms?

🞏 1-No enjoyment or pleasure

🞏 2-Little enjoyment or pleasure

🞏 3-Some enjoyment or pleasure

🞏 4-Much enjoyment or pleasure

🞏 5-Great enjoyment or pleasure

14. How often do you have painful orgasms?

🞏 5-Never

🞏 4-Rarely (once a month or less)

🞏 3-Sometimes (more than once a month, up to twice a week)

🞏 2-Often (more than twice a week)

🞏 1-Every day

### **CSFQ Scoring Instructions**

To score the CSFQ, sum the numerical value of each of the 14 items. For example, in Item 1, a response of “some enjoyment or pleasure” has a numerical value of 3, whereas a response of “much enjoyment or pleasure” has a numerical value of 4. Items 10 and 14 are reverse-scored: for example, on Item 10, a response of “never” has a numerical value of 5, whereas a response of “every day” has a value of 1.

To calculate the Total CSFQ score, add up the values of the responses for all 14 items. Items 10 & 14 are not included in the subscale scores, but are included in the total CSFQ score.

To calculate subscale scores, add up the values for only the items that correspond to a particular subscale.

Pleasure: Item 1

Desire/frequency: Items 2 + 3

Desire/interest: Items 4 + 5 + 6

Arousal/excitement: Items 7 + 8 + 9

Orgasm/completion: Items 11 + 12 + 13

Total CSFQ score: Items 1 through 14, or add all subscale scores plus items 10 & 14

**Interpretation of scores:**

Threshold scores were established by non-overlap of the 95% confidence intervals (CI) between patient’s and normal’s mean scores. CSFQ threshold scores indicating sexual dysfunction:

Men	Women

Pleasure	≤ 4	≤ 4

Desire/frequency	≤ 8	≤ 6

Desire/interest	≤ 11	≤ 9

Arousal/excitement	≤ 13	≤ 12

Orgasm/completion	≤ 13	≤ 11

Total score	≤ 47	≤ 41

Reproduced with permission of Anita H. Clayton, M.D.

### **Study Interview Questions**

Length: 30 minutes

Moderator: NA

Participant: patient affected by PF

**Introduction and Welcome (read to patient)**

- Thank you for participating in this interview
- The purpose of this interview is to gain a deeper understanding of the impact of participants’ PF diagnosis on their sexual activity, and generate new ideas not captured in the survey.
- Expectations of individual: open dialogue, please respond to questions and participate in discussion if you feel comfortable doing so.
- If you are uncomfortable at any point, you can choose to skip a question by saying “skip” and we will move on. If you would like to end the interview, you may indicate so and simply hang up the call.
- As the interviewer I will be asking a few questions. I may ask you to clarify a point but I will try to contribute to the discussion as little as possible.
- All data will be collected anonymously and confidentially. The session will be recorded. No names or identifying data will be used in data analysis.

**Opening question**

- What does sexual function mean to you? OR what made you choose to participate in this research?

**Questions**

- What does your sexual activity look like as a person living with interstitial lung disease?
- How has your PF diagnosis impacted your sex life? Have you had to change any behaviours? (If you’re comfortable, can you elaborate on how you have adapted)
- Do you think your sex life is affected more by your PF or other unrelated factors?
- How does oxygen affect your sex life (if using?)
- Have you found improvement or worsening in your sexual activity related to your PF treatments?
- What discussions, if any, have you had with your healthcare provider regarding sex or sexual activity. Tell me about that experience
- What resources or tools, if any, have you tried to address concerns with your sexual activity? Have you participated in pulmonary rehab, and were you exposed to these topics? How was it?
- What do you think limits discussing your sex life with your partner, friends, or healthcare workers?
- We have had less than expected uptake for this survey. Why do you think that is the case, and how do you think we can engage better with patients around this topic?
- What are your thoughts on healthcare providers addressing sex and sexual activity in people living with PF?
- Would you prefer your specialist or family doctor discuss your sexual activity with you? Would you prefer if it was a non-physician?
- Did the survey make you more interested in sexual activity and PF? If so, what other additional information do you want to know about sexual function and PF?

**Ending questions**

- Is there anything that wasn’t discussed you would like to bring up now?

**Conclusion**

- Thank you for your participation.

### **Qualitative codebook**

| **Parent Code: Sexual function understanding**   - **Patient understanding of sexual dysfunction** | | |
| --- | --- | --- |
| *Codes* | *Concept/description* | *Example* |
| Sexual function understanding | Illustrates the level of knowledge and understanding that patients have of sexual function as a concept | *“being able to perform you know, sexually, at least to be able to engage, you know pleasurably in in that experience.”* |
| **Parent code: Sex with PF**   - **Factors related to pulmonary fibrosis that impact the patient’s current or previous sex life** - **Changes that patients have made to their sex life since living with PF** | | |
| *Codes* | *Concept/description* | *Example* |
| **Sex with PF** | | |
| Current sex life | Patient’s description of current sexual activities | *“I think our sex life is like fairly active we have sex you know, weekly, probably.”* |
| Erectile dysfunction | Male patients’ experience with erectile dysfunction | *“I found that I no longer am able to get fully erect, since the fibrosis and I don't know if it's due to the fact I'm on so many medications… or if it's just something that's physically related, or change within my body, you know from disease.”* |
| Prioritizing sex life | Patient’s emphasis on continuing to be sexually active | *“It's probably one area of my life I try not to let affect, because everything else seems to be affected. But when I'm laying in bed, I can do me. I can be who I want to be. I don't have to worry about anything else you know, so it's that big, my time.”* |
| Adaptation | Patient’s experience in adjusting their sex life in context of PF | *“Definitely the sexual acts that we engage in have changed, I think, with the diagnosis and the progression”* |
| Desire | Patient’s desire for sex | *“[Symptoms of pulmonary fibrosis], well, it hasn't impacted my desire. you know, really at all.”* |
| Symptoms with sex | Patient’s experience during sexual activity | *“I think the fibrosis has really had a major a bigger impact on my sexual life, than I had anticipated. because it, you know, physically everything is harder to do. I have to pace myself on household chores and walking, things like that. So the energy consumed for sex would tire me out very quickly.”* |
| **Oxygen use** |  |  |
| **Feelings around oxygen** | |  |
| Oxygen shame | Feelings of shame around using oxygen during sex | *“How do you not notice the oxygen tubing on somebody's face, you know, or, you know, twisted up in, you know, in bed, or you, you know it's just. It can be awkward.”* |
| Self image with oxygen | Patient’s image of themselves with oxygen | *“I think it's had an effect in way, what is it? I hate it, I hate it on my face … It makes me almost feel like I'm separated from everybody else because of this thing that's on my face … You know it's changed how I smile. It's weird … I see myself, and like who is that? You know, and so it's just one more thing that just makes me a little self conscious.”* |
| **Sex with oxygen** | Patient’s experience of sexual activity while using oxygen | *“Now that I'm sitting here, I'm thinking I should probably turn it up during sex. But that's weird. It's like, Oh, hold on! I gotta get ready. Well, let me go turn up the juice. That's just awkward, you know, who's gonna do that?”* |
| **Parent code: Emotions**   - Various emotions and feelings that patients experience as it relates to their sex life | | |
| *Codes* | *Concept/description* | *Example* |
| Trust | Assured reliance on the character, ability, strength, or truth of another individual | *“Now she would be someone I would be quite comfortable talking about that basis with no problem at all. Whereas if it's one of the male doctors I will never see again, I'll be thinking … Why, does he want to know this?”* |
| Worry | Experience of concern or anxiety, typically for something impending or anticipated | *“I think if it were to get any worse, then I would certainly ask my primary care physician about it, and see what he would say”* |
| Priorities | Patient priorities based on emotional or physical needs | *“I was formally diagnosed in January of 2017, and at that point my attention turned completely to IPF and what to do about that”* |
| Embarrassment | Feeling of self-consciousness, discomfort, or shame | *“I think the only thing that would limit me talking to a professional, is if I felt I was making them uncomfortable, or embarrassed. Then I would probably hold back in the discussion.”* |
| Shame | Painful emotion caused by consciousness of guilt, embarrassment, or shortcoming, which may be physical | *“[My partner] says it doesn't bother him, he doesn't think about it, but I just can't imagine that that's a hundred percent true you know”* |
| Guilt | Belief of having done something wrong, or experiencing feeling bad, undeserving, or selfish | *“If I got the transplant, and there was something I had to give up. Is that really so bad?”* |
| Frustration | Feeling discouragement, anger, and annoyance due to unresolved problems, unfulfilled goals, desires, or needs. | *“Yeah, I hate that. I have this now. So it's definitely a struggle to live like a very healthy life.”* |
| **Parent code: Relationships**   - **The impact of PF and the patient’s sex life on their interpersonal relationships patients** - **Interest of patients in discussing their sex life with different interpersonal relationships** | | |
| *Codes* | *Concept/description* | *Example* |
| **Romantic Relationships** | | |
| LGBTQ relationships | Impact on lesbian, gay, bisexual, transgender, or queer relationships | *“I don't know how much information there would be that would be applicable to like, like, you know, a same sex relationship.”* |
| Partner concerns | Impact of pulmonary fibrosis on the patient’s romantic partner or spouse | *“..my wife is afraid of injuring me for one”* |
| Relational factors | Aspects of spouse/romantic partner relationship unrelated to pulmonary fibrosis | *“I think I have bigger relationship type issues that we've been working through that are unrelated to pulmonary fibrosis.”* |
| **Friend Relationships** | | |
| Friends sex discussions | Patient’s ability to discuss sex with non romantic friends | *“I feel that people of my generation fall into 2 categories, those who are comfortable talking about their sexual life and those who are not”* |
| **Healthcare Relationships** | | |
| Healthcare sex discussions | Previous discussions on sex with healthcare workers | *“…my general practitioner, I wouldn't have those conversations with them. Have I ever been asked trying to think back? I can't. I don't recall.”* |
| Interest in healthcare sex discussions | Patient’s interest in discussing sex with their healthcare workers | *“I think so, but not right away. You know I was scared to death when I got this diagnosis… So the last thing I'm thinking about is sex, you know at that point. And actually that was a long process for us to really get accustomed to, you know that it’s a life and death issue”* |
| Personal connection with healthcare worker | Patient’s personal connection with their healthcare workers or lack thereof | *“All of my doctors have seen the worst of me. The worst that my body can put me through. I'm very comfortable with all of them.“* |
| **Social support** |  |  |
| Social support | Other sources of social support for patient to discuss sex | *“The PF community post transplant is a very joyful, grateful community for the most part and I think that oftentimes they will short sell things that bring them great joy just because they feel like they've already been given their fair share.”* |
| **Parent code: Quality of Life**   - **Various consequences of PF that impact a patient’s quality of life, and therefore also impacts their sex life** | | |
| *Codes* | *Concept/description* | *Example* |
| Antifibrotic treatment | Effects of antifibrotic treatment (nintedanib, pirfenidone) on sexual activity and QoL | *“The treatment is a horrible little pill called Ofev, and 2 of the side effects are vomiting and diarrhea. So there will be occasions, and there have been, where you know, we may have felt in the mood for intimacy. The pill has decided that it's going to make me violently sick, or it's going to create a bout of diarrhea. So you have to put your sexual activity on the sidelines and wait until that side of life passes.”* |
| Comorbidities | Patient belief around how comorbidities affect their sexual activity | *“I also have heart disease… And I have always assumed that my erectile dysfunction had to do with my heart. And I have never actually had a diagnosis.”* |
| Pulmonary rehab | Patient experience of discussions about sex during pulmonary rehab programing | *“I did participate in pulmonary rehab. It was not a subject that was brought up in pulmonary rehab. It's just not a subject that's talked about really anywhere that I've seen.”* |
| Medication side effects | Patient symptoms related to medication side effects as it pertains to their sexual activity | *“I know medications play a large part in how I feel on any given day, even post transplant being. I think I'm on currently on 34 different pills a day and I know they play some part, and how the rest of my body is doing. But can I tie this directly to the medications? I don't know.”* |
| Symptom progression | Effect of PF symptom progression on sexual activity | *“Maybe further down the line, as my disease progresses more, I'll be like, you know, how can we engage in sex in a safe and happy way?”* |
| **Parent code: Research**   - Patient’s desire and experience in engaging in research - Patient’s own exploration to learn about PF and sexual function | | |
| *Codes* | *Concept/description* | *Example* |
| Importance of research | Patient perspective on why they have chosen to participate in this study, and their comments on the importance of this research area | *“I'm just a firm believer in research and you know, if somebody else can benefit from anything that I've had to say, that makes me happy.”* |
| Knowledge distribution | Patient perspective on sharing information around sexual activity and dysfunction in PF | *“I mean it has to be addressed. We can't keep sticking our heads in the sand and saying that it is not an issue.”* |
| Research engagement | Desire and values to participate in research, as well as being an active participant in finding information about PF | *“I just to try and help out. I don't, I don't have anything else. I was working, and now, I don't work anymore. So I have a lot of free time available.”* |
| Resources | Ability to find new research relating to PF | *“I feel like information in general, for people living with pulmonary fibrosis is hard to find, and it's very outdated on the Internet. so maybe you know, people aren't searching in the right places for this kind of research.”* |
